# Influence of liver attenuation on the severity of course COVID-19: a retrospective cohort study

**DOI:** 10.1101/2023.02.27.23286488

**Authors:** Yuliya F. Shumskaya, Anna P. Gonchar, Marina G. Mnatsakanyan, Ivan A. Blokhin, Roman V. Reshetnikov, Yuriy A. Vasilev

**Affiliations:** Research and Practical Clinical Center for Diagnostics and Telemedicine Technologies of the Moscow Health Care Department, 24, Petrovka str., bldg. 1, Moscow, 127051, Russia; I.M. Sechenov First Moscow State Medical University, Ministry of Health of Russia (Sechenov University), 8, Trubetskaya str., bldg. 2, Moscow, 119991, Russia

**Keywords:** COVID-19, SARS-CoV-2, liver attenuation, computed tomography, automatic segmentation

## Abstract

**Introduction:** There is no unequivocal opinion concerning the influence of decreased liver attenuation on the COVID-19 severity, but its widespread occurrence among these patients has been shown. It is still debatable whether decreased liver attenuation is an independent risk factor for the severe course of CVID-19.

**Study objective:** To assess the prognostic value of liver attenuation on CT scan in patients with COVID-19.

**Material and methods:** A retrospective cohort study. Data of COVID-19 outpatients were analyzed. Inclusion criteria: two chest CT scans, alanine aminotransferase (ALT) and aspartate aminotransferase (AST) levels in blood, and polymerase chain reaction results to verify SARS-CoV-2. Subjects were categorized into four comparison groups depending on the severity of lung involvement. Liver attenuation was analyzed by automatic segmentation, where the values less than 40 HU were considered pathological.

**Results:** Data from 499 subjects were included. The groups differed in age and the level of liver attenuation on both CT scans. No correlation between ALT, AST and changes in liver attenuation was found. On follow-up CT, low liver attenuation was observed in males (odds ratio (OR) 2.79 (95% CI 1.42–5.47), *p-value* = 0.003) and in patients with a baseline reduced liver attenuation (OR 60.59 (95% CI 30.51–120.33), *p-value* < 0.001). Age over 60 years was associated with the development of lung lesions (OR 1.04 (95% CI 1.02–1.06) for extent of lung injury < 25%, OR 1.08 (95% CI 1.05–1.11) for 25–50%, OR 1.1 (95% CI 1.06–1.15) for 25– 50%, *p-value* < 0.001). Low liver attenuation on the baseline CT scan increased the odds of severe lung injury (OR 6.9 (95% CI 2.06–23.07), *p-value* = 0.002).

**Conclusion:** In COVID-19, patients with low liver attenuation are more likely to develop severe lung damage.

## Introduction

The new coronavirus infection disease (COVID-19), initially registered in the city of Wuhan in the People’s Republic of China in December 2019, caused a global pandemic. The mortality rates among the affected patients range from 10% to 26% [1]. In view of the above figures, accurate risk assessment for each patient is believed to be of vital importance. To achieve this, we must develop an understanding of the factors that predispose severe COVID-19. The available studies provide insight into the following risk factors that predispose severe course of the new coronavirus infection: age over 60 years, excess body mass, male gender, essential hypertension, and type 2 diabetes mellitus [2]. The studies also consider chronic obstructive pulmonary disease [3], bronchial asthma [4], kidney disease [5], and liver disease [6] as the possible risk factors.

There is conflicting evidence regarding the impact of metabolic-associated fatty liver disease (MAFLD) on the COVID-19 severity. Some studies found no relationship between the two [7], while others show that MAFLD has a negative impact on COVID-19 progression [8]. The above-mentioned studies relied on decreased liver attenuation of less than 40 HU on computed tomography (CT) scans as a diagnostic criterion of MAFLD. Currently, there is no studies that assess the liver status in patients with COVID-19 both before and during the development of SARS-CoV-2-associated lung injury – a correlation that sparked a particular interest in this study.

The goal of this study was to assess the prognostic value of liver attenuation on CT scans in patients with COVID-19.

## Material and methods

Study type: A retrospective cohort study carried out using the “Standards for reporting original research for observational studies: STROBE” guidelines. Given the retrospective design of this study, the subjects were not required to fill in informed consent froms. This study was approved by the independent ethics committee of the Moscow Regional Branch of the Russian Society of Roentgenologists and Radiologists (MRB RSRR).

For this study, we analyzed data from outpatients who underwent chest CT during screening for a lung injury associated with COVID-19 that took place from January to July 2020. A total of 139,590 patients were included in the study database. *Inclusion criteria:* two chest CT scans; measured alanine aminotransferase (ALT) and aspartate aminotransferase (AST) levels in blood; results of a polymerase chain reaction (PCR) throat swab to verify SARS-CoV-2. *Exclusion criteria:* age below 18 years; pregnancy; availability of only one chest CT study; no data on either the ALT or AST levels; no PCR throat swab results for SARS-CoV-2 verification; lung injury features corresponding to that associated with SARS-CoV-2 on baseline CT; malignant mass in the liver evidenced by a change in its density; iron-rich inclusions in the liver; incorrect segmentation of the liver; errors in the automatic analysis of the liver attenuation.

The study presents four comparison groups. The distribution criteria are presented in *Table 1*

**Table 1.**
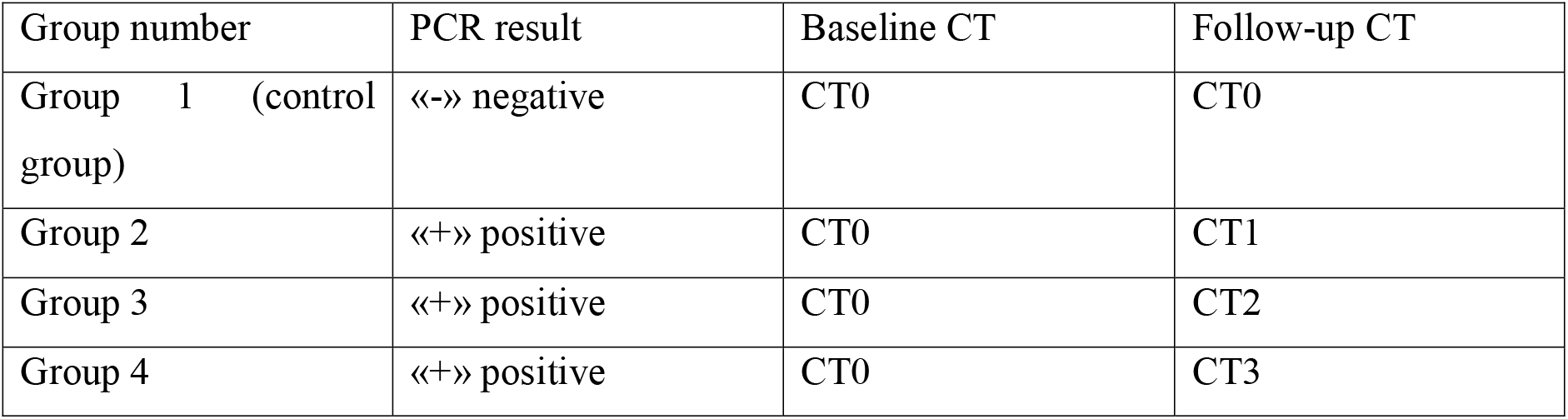
Inclusion criteria for the comparison groups

| Group number | PCR result | Baseline CT | Follow-up CT |
| --- | --- | --- | --- |
| Group 1 (control group) | «-» negative | CT0 | CT0 |
| Group 2 | «+» positive | CT0 | CT1 |
| Group 3 | «+» positive | CT0 | CT2 |
| Group 4 | «+» positive | CT0 | CT3 |

The attenuation of less than 40HU was considered a sign of low liver density. Elevated hepatic transaminase values were considered as ALT> 40 U/L, AST> 41 U/L.

All studies were performed on Toshiba Aquilion 64, Toshiba Aquilion CXL, and General Electric HiSpeed CT scanners. The chest examination was performed according to the standard protocol: tube voltage – 120 kV; automatic current modulation depending on the scout view; scanning direction – from the diaphragm to the apex of the lungs; FOV – 350 mm; slice thickness ≤ 1 mm; kernel – soft tissue. The scanning was performed with breath-holding at inspiration depth. Using the CT 0–4 empirical visual scale, radiology specialists of outpatient CT centers with 8 to 22 years of experience performed primary readings of the chest CT images available in the Unified Radiological Information Service. The automatic assessment of the liver attenuation was performed using the method that involved automatic segmentation of the liver based on the correlation between its initial shape and the embedded templates, followed by the evaluation of the average parenchymal density in the ROI [9].

The data analysis was performed using the R 4.2.0 software and coding language. All subjects with valid CT liver density data were included in the data analysis. The following values were analyzed: liver attenuation at two time points; ALT and AST levels. The normality assessment was performed using the Shapiro-Wilk test. To present quantitative variables we used the following measurements: mean value and standard deviation for normally distributed variables; median and interquartile range, otherwise. To assess the correlation between the variables, we used the Pearson coefficient (for normally distributed data) or the Spearman coefficient (otherwise). Comparison of the variable values across the four groups was carried out using analysis of variance (Fisher ANOVA for normally distributed data, Kruskal-Wallis ANOVA – otherwise). For pairwise comparison, we performed a post-hoc analysis (ANOVA post-hoc). Analysis of variance was carried out using the ggstatsplot package. To reduce the false discovery rate for the null hypothesis, the Benjamini-Hochberg procedure was used. We used multivariate regression analysis to assess the impact of the putative risk factors (age, gender, reduced liver attenuation on baseline CT, elevated ALT, and/or AST) on the severity of lung injury over time.

## Results

For this study, we used the examination results of 515 subjects out of a total of 139,590 available (Fig. 1). Sample composition: 198 men (38.4%) and 317 women (61.6%); median age – 47 years (from 18 to 91 years). The average gap between the two CT examinations in groups 1 and 2 was 14 days; group 3 – 7 days; group 4 – 14 days.

**Fig. 1.**
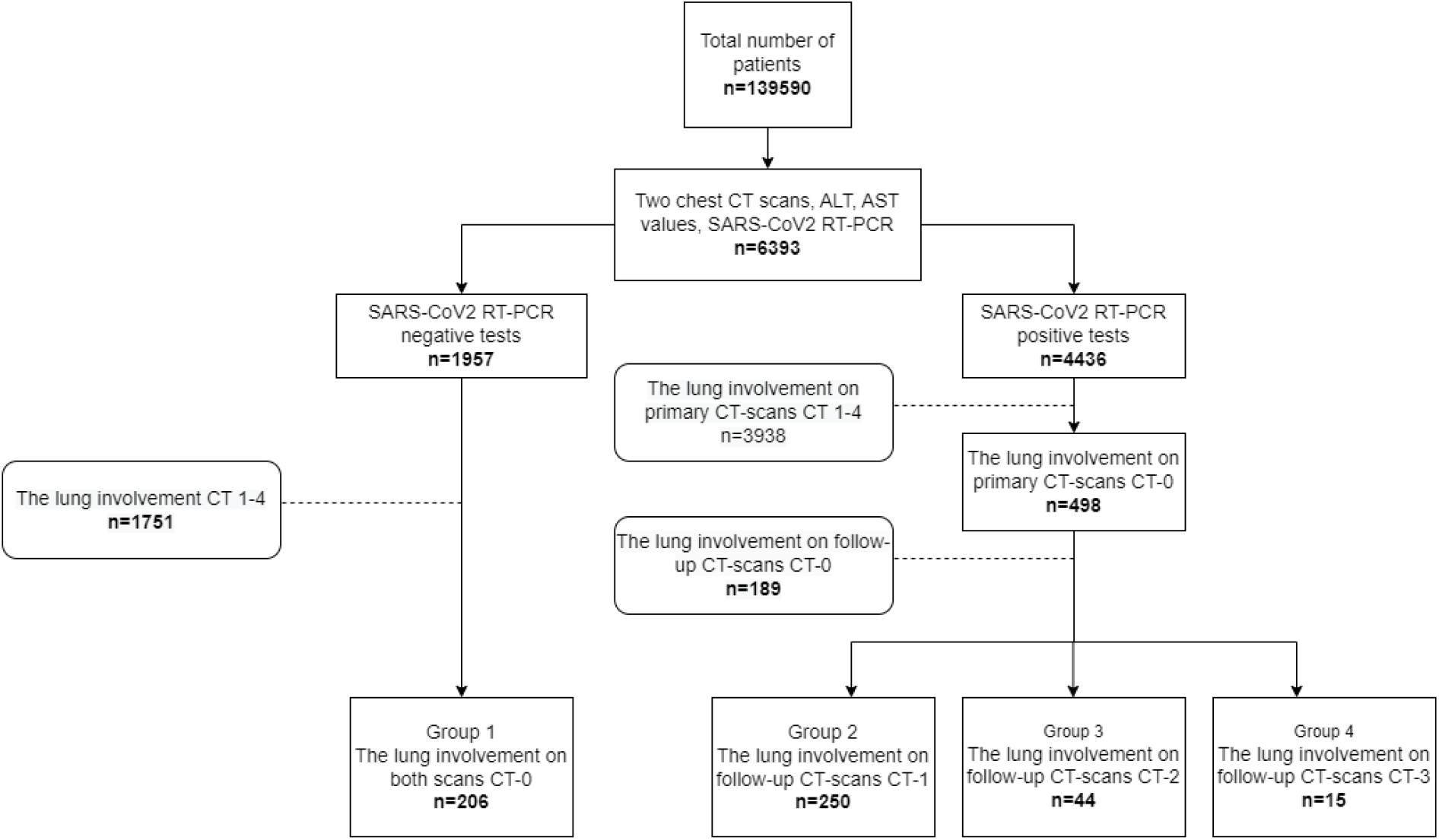
Sampling workflow

Due to errors in the segmentation algorithm, we excluded 16 subjects from the analysis and then built the comparison groups. The features of the comparison groups are presented in *Table 2*.

**Table 2.** Clinical characteristics of the study groups

| Parameter | Group 1 | Group 2 | Group 3 | Group 4 |
| --- | --- | --- | --- | --- |
| Total number of patients, *n* | 206 | 250 | 44 | 15 |
| Included patients, *n* | 194 | 246 | 44 | 15 |
| Female, *n* (%) | 126 (64,9) | 154 (62,6) | 27 (61,4) | 6 (40) |
| Male, *n* (%) | 68 (35,1) | 92 (37,4) | 17 (36,6) | 9 (60) |
| Age | 39,5 [32; 50] | 49,5 [38; 59] | 55,8±12,1 | 60,6±15,3 |
| ALT, u/l | 20,1 [15,1; 30,9] | 22 [15; 34,15] | 28,75 [17,85; 44,9] | 19 [13,8; 29,6] |
| AST, u/l | 22,9 [19; 27,9] | 24,1 [19,9; 30,35] | 32,1 [24,35; 45,45] | 23,9 [22,45; 30,4] |
| Liver CT-attenuation on baseline CT, HU | 53,5 [44,3; 58,6] | 47,5 [39,95; 54,9] | 45,7±11,26 | 38,94 [35,1; 45,2] |
| Liver CT-attenuation on follow-up CT, HU | 52,4 [44,75; 58,95] | 48 [41,4; 54,8] | 46,7 [40,1; 53] | 37,56±11 |
| **Liver CT-attenuation <40 HU on the baseline CT** |  |  |  |  |
| Total number of patients, *n* (%) | 31 (16,0) | 62 (25,2) | 13 (29,5) | 10 (66,7) |
| Female, *n* (%) | 16 (51,6) | 28 (45,2) | 6 (46,2) | 5 (50) |
| Male, *n* (%) | 15 (48,4) | 34 (54,8) | 7 (53,8) | 5 (50) |
| Age | 45±11,3 | 52±11,6 | 52,1±11,8 | 66,7±13,5 |
| ALT, u/l | 33 [23; 57,4] | 30,25 [23,6; 47,5] | 39,4 [30,5; 44,6] | 25,2±17 |
| AST, u/l | 27 [23; 43] | 29,3 [22,4; 38,9] | 37 [26; 45,7] | 28,7±11,9 |
| **Liver CT-attenuation <40 HU on the follow-up CT** |  |  |  |  |
| Total number of patients, *n* (%) | 27 (13,9) | 53 (21,5) | 11 (25) | 7 (46,7) |
| Female, *n* (%) | 14 (51,9) | 20 (37,7) | 5 (45,5) | 1 (14,3) |
| Male, *n* (%) | 13 (48,2) | 33 (63,3) | 6 (54,6) | 6 (85,7) |
| Age | 42±11,8 | 52,6±10,8 | 58,6±11,4 | 54,7±13,2 |
| ALT, u/l | 29 [21,15; 51,45] | 30 [22,8; 43] | 37,2±12,8 | 28,5±11,8 |
| AST, u/l | 25 [21; 32,55] | 28,1 [22,2; 36,5] | 38,9±15,1 | 24,9±6,3 |

The occurrence of low liver density at the onset of COVID-19 was 23.2%, while during the peak of the outbreak, it was 19.6%.

When evaluating the relationship between ALT, AST, and changes in liver density over time, no positive correlation was observed (Spearman correlation coefficient: for ALT – 0.027, *p-value* = 0.54; for AST – 0.004, *p-value* = 0.9).

Comparison of the age groups revealed statistically significant differences across all the groups (*p-value* < 0.001), except for groups 3 and 4 (*p-value* = 0.47).

Liver densities on baseline CT (*Fig. 2*) also vary significantly across all groups (*p-value* < 0.001), except for groups 2 and 3 which demonstrated no statistically significant difference (*p-value* = 0.63).

**Fig. 2.**
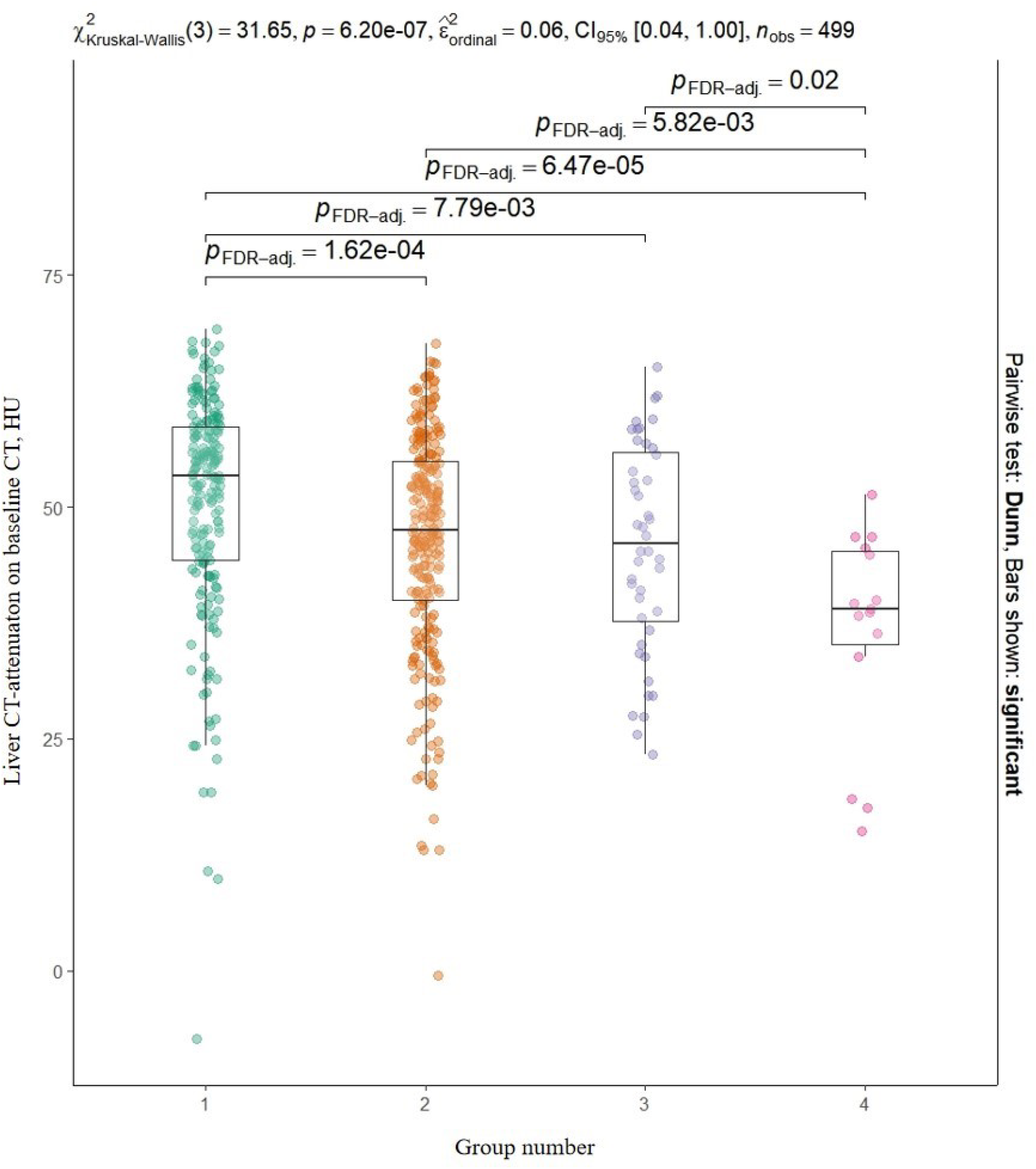
Analysis of variance for the comparison groups by liver density on baseline CT with post-hoc analysis

Liver density values on CT (*Fig. 3*) also do not differ over time across groups 2 and 3 (*p-value* = 0.34) and groups 3 and 4 (*p-value* = 0.05), as opposed to all the remaining groups (*p-value* < 0.001).

**Fig. 3.**
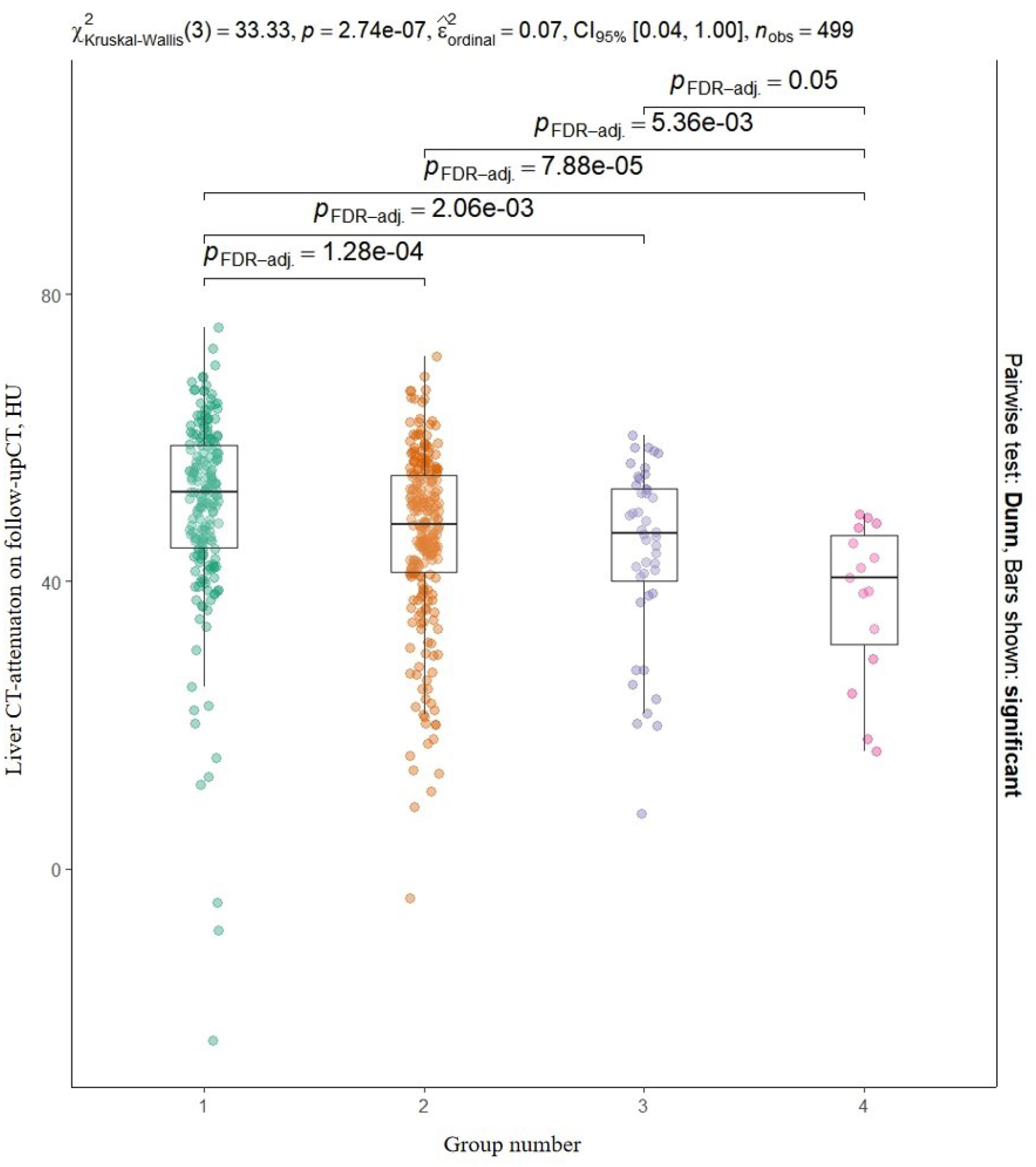
Analysis of variance for the comparison groups by liver density on follow-up CT with post-hoc analysis

When comparing the changes in liver attenuation over time, no significant difference was observed across all the groups (Kruskal-Wallis test: *p-value* = 0.54).

For all three comparison groups, age over 60 years is associated with the development of lung injury. For >25% pulmonary involvement (CT1) in group 2, OR was 1.04 (95% CI 1.02-1.06), *p-value* < 0.001. For 25-50% pulmonary involvement (CT2) in group 3, OR was 1.08 (95% CI 1.05-1.11), *p-value* < 0.001. For acute pulmonary involvement (CT3) in group 4, OR was 1.1 (95% CI 1.06-1.15), *p-value* < 0.001.

In addition, patients with higher AST were more likely to develop 25%–50% pulmonary involvement on a follow-up chest CT (CT2) – OR 8.05 (95% CI 2.7–23.98), *p-value* < 0.001.

Patients with low liver density on baseline chest CT had greater odds of acute pulmonary involvement on a follow-up chest CT– OR 6.9 (95% CI 2.06–23.07), *p-value* = 0.002.

## Discussion

The study found that patients with diagnosed SARS-CoV-2 infection with low liver density on baseline chest CT are more likely to develop acute lung injury over time (OR 6.9; *p-value* = 0.002). At the same time, elevated ALT and AST are not associated with low liver density.

Previous papers explained the low liver density during COVID-19 by the damage attributed to various factors: the virus itself, the administered medications, etc. [10, 11]. This allowed us to suggest, that the changes in liver density must correlate with an increase in the degree of lung involvement over time. However, our data did not confirm such a relationship.

We have determined that low liver density on baseline CT correlates with much higher risks of developing acute lung involvement over time. It is known that an increase in lung tissue damage on CT according to the CT0–4 scale has a direct correlation with an increased risk of death. Thus, the risks for patients with CT4 lung involvement are 3 times greater than for those with CT0 [12]. Since the same liver changes are also seen in the diseases caused by metabolic syndrome, it was possible to draw a link between metabolic co-morbidities and complications caused by COVID-19. The existence of such a relationship is confirmed by several authors [13].

In their study, P. Lei et al. determined that a quarter of all CT studies of the upper abdominal cavity in patients with COVID-19 show a decreased liver attenuation. In our study, the proportion of patients with decreased liver attenuation did not exceed 1/5 of the sample. At the same time, the likelihood of low liver density grows significantly as COVID-19 advances to more severe stages, which does not contradict the literature data [14]. However, these studies either deny the presence of previous liver changes or provide no such information.

E. Guler et al. found, that patients with COVID-19 whose chest CT images show lung injury getting worse over time have a greater chance to develop low liver density [15]. However, our data show no statistically significant differences between the rate of liver density changes across the groups and the development of pneumonia of varying degrees. This discrepancy may be attributed to a notable difference in the sample sizes (27 vs. 499), as well as to the method we used to determine low liver density based on the ratio of liver density to spleen density.

In our study, it was determined that the probability of low liver density in males with severe COVID-19 is higher compared to females, which was also observed in other studies [2]. Taking the above data into consideration, it became possible to single out a separate risk group among the male population.

We noted the absence of a correlation between liver density values over time and elevated ALT and AST. Researchers show that an increase in transaminase is associated with an increased risk of severe COVID-19 and the related mortality [16]. On the other hand, in their study, Y. Zhang et al. found no relationship between elevated transaminase and SARS-CoV-2, which is confirmed by our data [17]. In addition, some authors question the clinical significance of changes in the transaminase level [18].

Our study provides evidence for the importance of identifying signs of metabolic syndrome in patients with COVID-19, especially decreased liver attenuation on chest CT. It must be mentioned, that metabolic-associated fatty liver disease is observed in most patients with obesity, which is also associated with more severe COVID-19 [19].

Decreased liver attenuation observed on baseline CT may become a valuable and most importantly a reliable predictor of a widespread lung involvement in COVID-19 and more severe course of the disease. Our data and the findings of the above studies evidence the need to single out patients at higher risk of complications in COVID-19 for accurate risk stratification, more thorough patient management and medication choice.

## Limitations

Our study has several limitations. The use of the CT0–4 scale to grade the lung involvement along with a visual semi-quantitative assessment of lung parenchymal damage limits the generalizability of the results obtained at the international level. A strict 40 HU liver attenuation threshold also imposed its limitations on the obtained results. More flexible thresholds would have led to changes in the data, i.e., an increase in the number of patients with decreased liver attenuation. We also consider smaller sample sizes of groups 3 and 4 relative to groups 1 and 2 as limitations.

## Data Availability

All data produced in the present work are contained in the manuscript

